# Attitudes towards new tuberculosis vaccines among adults, adolescents and their caregivers in southern Mozambique, 2024

**DOI:** 10.64898/2026.03.30.26349798

**Authors:** Agostinho Lima, Ilse Campos, Doyoon Kim, Machi Shiiba, Lisa Marie Cranmer, Sozinho Acácio, Alberto L. García-Basteiro, Lavanya Vasudevan, Kristin Nelson

## Abstract

New tuberculosis (TB) vaccines for adults and adolescents could transform TB prevention programs, but their impact depends on successful implementation. We investigated willingness to be vaccinated with a new TB vaccine in a high HIV and TB burden setting in southern Mozambique in 2024 using a mixed methods approach involving a cross-sectional survey and concurrent in-depth interviews. In 151 surveys and 23 interviews, we found that willingness to receive a new TB vaccine among adults and adolescents was 77% (148/192) overall. In multivariable analysis, adolescents were more willing to receive a new TB vaccine than adults even when adjusting for other factors which may influence vaccination decisions (adjusted OR: 5.6, 95% CI: 1.7-17.7). Personal experience with TB and greater knowledge of the disease was also linked with willingness to be vaccinated. Qualitative findings reinforced quantitative findings, further clarifying that even among those who expressed hesitancy, a safe and effective TB vaccine endorsed by healthcare workers, government agencies, and community leaders would likely have high uptake. Our findings are specific to southern Mozambique and can shape vaccine introduction efforts after a TB vaccine is licensed and approved for use in this age group.

## Background

More than 10 million people develop tuberculosis (TB) every year.^1^ Progress towards global goals of reduced morbidity and mortality from TB has been slow for the past several decades and was significantly delayed by the recent COVID-19 pandemic. However, new TB vaccines currently under development for adults and adolescents could transform TB prevention programs in high-burden settings.^2^

The impact of a new efficacious TB vaccine will be dependent on widespread population coverage in high TB burden settings. Ensuring high demand for a new vaccine is particularly a concern against the backdrop of a global crisis in vaccine confidence, which was intensified by the COVID-19 pandemic. In a study of 23 countries conducted in 2023, nearly a quarter of survey respondents were less willing than before the COVID-19 pandemic to receive routine immunizations.^3^ If not directly addressed, sustained skepticism towards vaccines represents a threat to the effectiveness of future TB vaccine programs. Attitudes towards new vaccines vary. A multi-country study of malaria vaccine acceptance completed prior to introduction reported wide ranges of willingness to accept malaria vaccine by setting, with acceptance as low as 30% reported in some studies but as high as 99% in other studies which focused on malaria-endemic areas.^4^ Another study reported that over 90% of adolescent girls in Mozambique would be willing to receive a vaccine against human papillomavirus (HPV) if it was available to them.^5^ Importantly, attitudes towards a vaccine can be also shaped by the perceived severity of the target disease and a person’s risk of being affected.^6^ Although coverage for the Bacille Calmette-Guerin (BCG) vaccine, which is given to infants in most high TB burden countries, is high (83-94% in 2024 across WHO regions)^7^, these attitudes may not translate to attitudes towards a new TB vaccine for adults and adolescents. Understanding the demand for new TB vaccines among people who would be eligible for them and, more broadly, exploring how attitudes and perceptions about vaccines generally may shape the decision to receive a TB vaccine, can set expectations for initial demand and support planning for communication and outreach strategies to maximize uptake.

Understanding attitudes towards new TB vaccines will be essential in countries with the highest TB burden, where vaccines stand to have the greatest health impact. We investigated willingness to be vaccinated with a new TB vaccine in a high HIV and TB burden setting in southern Mozambique in 2024 using a mixed methods approach involving a cross-sectional survey and concurrent in-depth interviews. Our mixed methods approach allowed us to both explore statistical associations between individual attributes and willingness to vaccinate as well as characterize emergent, and possibly unanticipated, attitudes and perceptions about new TB vaccines. We present in this analysis the integrated findings of the quantitative and qualitative data from this study.

## Methods

We followed the Good Reporting of A Mixed Methods Study (GRAMMS) checklist for the reporting of this study.^8^ (Supplement Section A)

### Study Population and data collection

The study was conducted in Manhiça district, southern Mozambique, from February to July 2024. In 2024, the estimated incidence of TB in Mozambique was 361 per 100,000. The TB epidemic in Mozambique is driven by HIV, with approximately 25% of incident TB occurring among people living with HIV.^1,9,10^ Participants were recruited from the community through household sampling facilitated by the demographic surveillance system in Mahniça^11^, or from the TB outpatient clinic at Manhiça District Hospital, which serves patients receiving treatment for TB. In the community, the study team visited households to offer participation to all eligible household members. Patients attending the TB clinic were approached for participation at the end of their clinical visit.

Eligible participants included adults (aged 18 years or older), adolescents (aged 9 to 17 years), and parents or caregivers of adolescents (aged 18 or older). Written consent was obtained from all participants over the age of 17. For participants aged 17 and under, written assent and parental consent were obtained. There were no exclusion criteria for participation.

In surveys, we collected demographic information, participants’ prior vaccination experiences, sources of information and beliefs about vaccination. We provided brief, general information about new TB vaccines under development and asked participants whether they would be willing to receive a novel TB vaccine. Survey items and scales were developed using validated survey tools based on the domains of the Behavioral and Social Drivers of Vaccination (BeSD) framework for vaccine hesitancy.^12^ Surveys were conducted in the participants’ preferred local language by trained field staff.

During the survey, all respondents were offered the opportunity to participate in an in-depth interview. From those who agreed, we purposively selected for interview a group who would represent the groups included and range of perspectives on vaccination expressed in the survey (willing and unwilling to receive a TB vaccine, TB patients and non-TB patients, those reporting past negative vaccine experiences and those not). Interviews were conducted in the participants’ preferred local language (Portuguese or Changana) by trained field staff, then transcribed and translated into English prior to analysis.

We aimed to recruit 250 participants for surveys and, among those, 24 for interviews. The sample size for the survey was based on our objective of estimating the proportion of persons willing to be vaccinated (expected at 80%) with a confidence interval width of 10% and allowed for multivariable analysis with up to 10 variables with several levels each. Sample size for the interviews was chosen to achieve code saturation (the point when no additional issues are identified and the codebook begins to stabilize) rather than meaning saturation (the point when issues are fully understood and no further insights can be found).^13,14^

### Mixed methods approach

We used a convergent mixed methods design, in which quantitative and qualitative data are collected concurrently with equal emphasis placed on both methods.^15^ Our quantitative analysis made use of participant responses to survey items about willingness to vaccinate with a new TB vaccine and attitudes, experiences, and perceptions about vaccination and TB that could be related to this decision. The qualitative analysis made use of participant interviews, which were meant to explore the perceptions and attitudes driving survey responses. The instrument joint display is presented in supplementary files (Supplement Section B).

### Quantitative data analysis

We calculated the proportion of individuals willing to receive a new TB vaccine overall. For the univariable and multivariable analysis, we conservatively classified ‘yes’ responses as willing to receive a vaccine; ‘no’ and ‘unsure’ responses were classified as unwilling to receive a vaccine.

We constructed variables describing past experiences and knowledge of TB from combinations of survey questions. We assessed participants’ TB knowledge level based on their agreement with three statements, to which they could have answered ‘Yes’, ‘No’ or ‘Unsure’: (1) “TB is a serious disease”; (2) “There is treatment available for tuberculosis”; and (3) “It is possible to die from tuberculosis”. An affirmative response to all three questions was considered a high level of TB knowledge, to two questions was considered a moderate level, and to one or zero questions was considered a low level. We assessed participants’ perception of the risk of TB based on their level of agreement with two statements: (1) “Tuberculosis is a common disease in my community” and (2) “I can easily catch tuberculosis.” Participants rated their answers on a scale from 1 (completely agree) to 6 (completely disagree). We grouped ratings 1–3 as “Agree” and 4–6 as “Disagree” for each question. Agreement with both statements was classified as a high level of perceived risk, with one statement was classified as a moderate level, and with no statement was classified as a low level of perceived risk.

We conducted univariate analyses and multivariable logistic regression to assess associations between willingness to be vaccinated with a novel TB vaccine and participant characteristics, including age group (adolescent, adult), previous vaccination experiences, and knowledge of TB. We considered variables reflecting previous vaccination experiences and knowledge of TB as potential causes of willingness to receive a vaccine. We used p = 0.05 as a threshold of statistical significance but report p-values and 95% confidence intervals for all odds ratios. All analyses were conducted using R software.

### Qualitative data analysis

We conducted thematic analysis on transcribed interviews from adults and caretakers of adolescents. Due to considerably lower saturation, interviews of adolescents were primarily used to triangulate findings from the survey and to confirm adult and caregiver findings. For initial data exploration, we selected a subset of transcripts (n = 5) with the richest responses. Using these five transcripts, we developed the codebook using an open coding approach based on the frequency of intersections of codes within text segments. The same set of codes was applied to adolescent transcripts.

All relevant codes under each theme and their associated segments were transcribed to generate summaries of each participant’s expression of the theme. These segment matrices were used to further elucidate patterns within and across participants and to elaborate on the identified themes.

Through this process, we developed nine main themes, classified into the domains of the BeSD framework.^16^ (See Supplement section D for detailed code descriptions). Themes and sub-themes, as well as illustrative quotes, were reviewed and approved by the field team in Mozambique. A 10% sample of transcripts (n=3) were coded by a second team member to assess inter-coder agreement. The frequency of the same code occurrences between the two coders was compared, which was determined to be adequate (>80%). All qualitative analysis was performed using MAXQDA 24 software. Illustrative quotes from the interviews are presented to support thematic findings.

Integration of qualitative and quantitative finding were performed using a joint display which was drafted by one team member and critically reviewed by another. Field staff reviewed the joint display to ensure fidelity to their experiences conducting surveys and interviews with participants.

We have made available an abbreviated dataset and the scripts used for the descriptive and multivariable analysis of survey data on GitHub https://github.com/kbratnelson/tbvax-willingness-moz). Qualitative data is available upon reasonable request subject to ethical approval and data privacy restrictions.

This study was approved by ethical and scientific committees at Centro de Investigação em Saúde da Manhiça (CISM), Manhiça, Mozambique (CIBS-CISM/050/2023 and CCI/036/AGO/2023) and Emory University (study #00006480).

## Results

### Survey findings

We enrolled 151 adults (median age 34, 61% female), 41 adolescents (median age 14, 66% female), and 49 caregivers (median age 30, 90% female). Most respondents (n = 163, 68%) were female. About half of participants (n = 134, 56%) had been previously diagnosed with TB. (Table 1) Among adults and adolescents, willingness to receive a new TB vaccine was 77% (148/192) overall. Willingness was higher among adolescents (88% ‘Yes’ responses) than adults or caregivers (74% ‘Yes’ responses) (one-sided p value = 0.045). Eight percent (8%) of participants responded that they would not receive a new TB vaccine (a ‘No’ response). (Table 1) Of note, adult men were less willing to be vaccinated than women (20% of men responded ‘No’ as compared to 4% of women, one-sided p value = 0.005). Adult women were more likely to express uncertainty (7% of men responded ‘Maybe’ as compared to 22% of women, one-sided p value = 0.011) in their intention to be vaccinated.

**Table 1:**
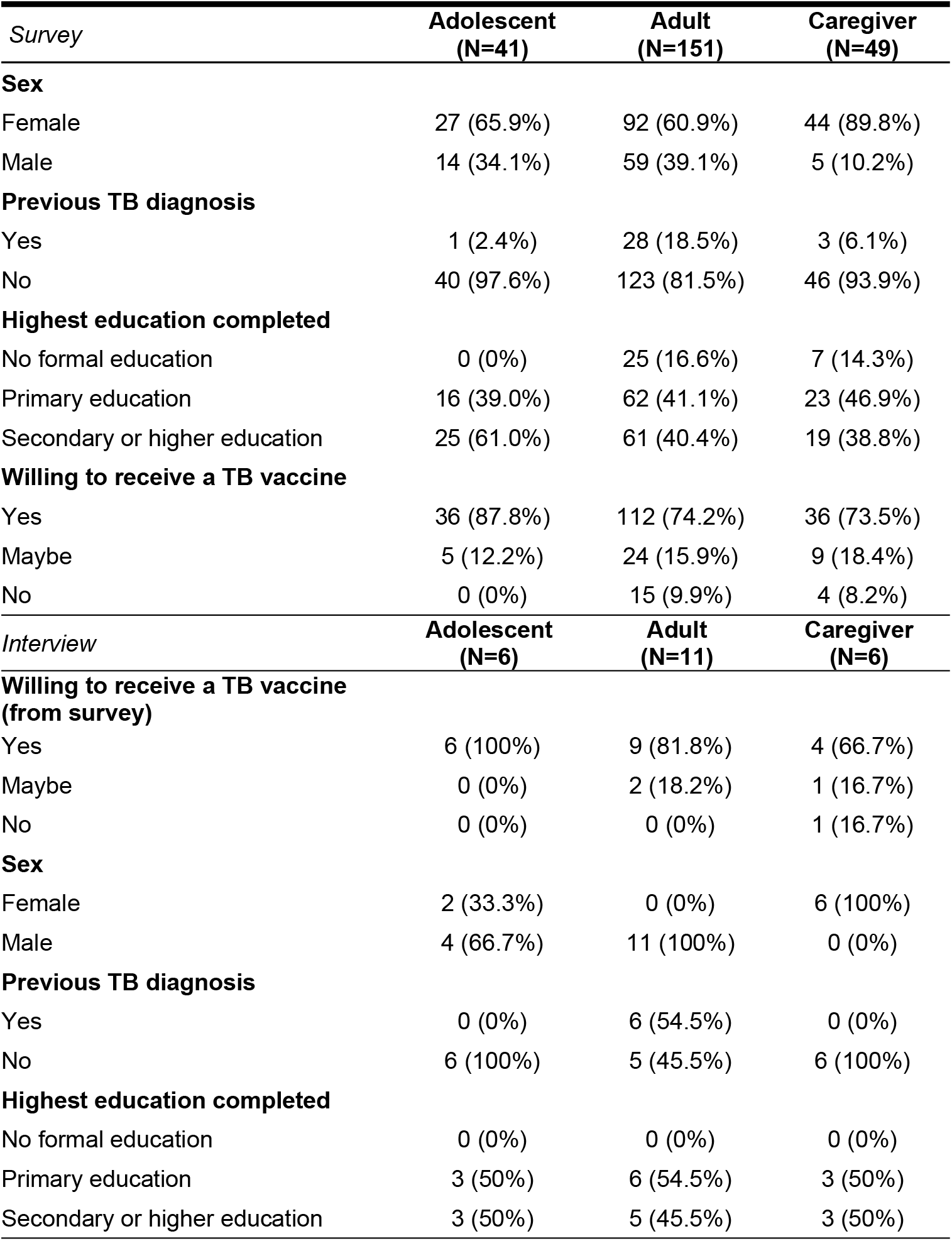
Participant demographics.

| *Survey* | **Adolescent**<br>**(N=41)** | **Adult**<br>**(N=151)** | **Caregiver**<br>**(N=49)** |
| --- | --- | --- | --- |
| **Sex** |  |  |  |
| Female | 27 (65.9%) | 92 (60.9%) | 44 (89.8%) |
| Male | 14 (34.1%) | 59 (39.1%) | 5 (10.2%) |
| **Previous TB diagnosis** |  |  |  |
| Yes | 1 (2.4%) | 28 (18.5%) | 3 (6.1%) |
| No | 40 (97.6%) | 123 (81.5%) | 46 (93.9%) |
| **Highest education completed** |  |  |  |
| No formal education | 0 (0%) | 25 (16.6%) | 7 (14.3%) |
| Primary education | 16 (39.0%) | 62 (41.1%) | 23 (46.9%) |
| Secondary or higher education | 25 (61.0%) | 61 (40.4%) | 19 (38.8%) |
| **Willing to receive a TB vaccine** |  |  |  |
| Yes | 36 (87.8%) | 112 (74.2%) | 36 (73.5%) |
| Maybe | 5 (12.2%) | 24 (15.9%) | 9 (18.4%) |
| No | 0 (0%) | 15 (9.9%) | 4 (8.2%) |
| *Interview* | **Adolescent**<br>**(N=6)** | **Adult**<br>**(N=11)** | **Caregiver**<br>**(N=6)** |
| **Willing to receive a TB vaccine**<br>**(from survey)** |  |  |  |
| Yes | 6 (100%) | 9 (81.8%) | 4 (66.7%) |
| Maybe | 0 (0%) | 2 (18.2%) | 1 (16.7%) |
| No | 0 (0%) | 0 (0%) | 1 (16.7%) |
| **Sex** |  |  |  |
| Female | 2 (33.3%) | 0 (0%) | 6 (100%) |
| Male | 4 (66.7%) | 11 (100%) | 0 (0%) |
| **Previous TB diagnosis** |  |  |  |
| Yes | 0 (0%) | 6 (54.5%) | 0 (0%) |
| No | 6 (100%) | 5 (45.5%) | 6 (100%) |
| **Highest education completed** |  |  |  |
| No formal education | 0 (0%) | 0 (0%) | 0 (0%) |
| Primary education | 3 (50%) | 6 (54.5%) | 3 (50%) |
| Secondary or higher education | 3 (50%) | 5 (45.5%) | 3 (50%) |

Of these participants, 23 participated in in-depth interviews. Most interview participants (n=15, 65%) were men, though all caregivers who participated were women. Four (4) interview participants out of 23 (17%) responded ‘No’ or ‘Maybe’ when asked if they would receive a new TB vaccine.

In a multivariable logistic model including demographic factors, previous vaccination experiences, and knowledge/experience of TB, we found that the strongest determinant of willingness to vaccinate was age: adolescents were nearly six times more willing to receive a new TB vaccine as adults (adjusted OR: 5.6, 95% CI: 1.9-19.6). Participants who reported previous issues accessing vaccines were more than three times as willing to receive a new TB vaccine than those who did not report previous access issues (adjusted OR: 3.3, 95% CI: 1.3-8.9). Although not significant at p = 0.05, we also found that participants reporting personal experience with TB (a friend or family member with the disease) (adjusted OR: 2.4, 95% CI: 0.8-7.9) and with a higher knowledge of TB (adjusted OR 2.6, 95% CI: 0.9-7.5) were more willing to receive a new TB vaccine. There were no missing responses for the variables used in the model. (Table 2)

**Table 2.**
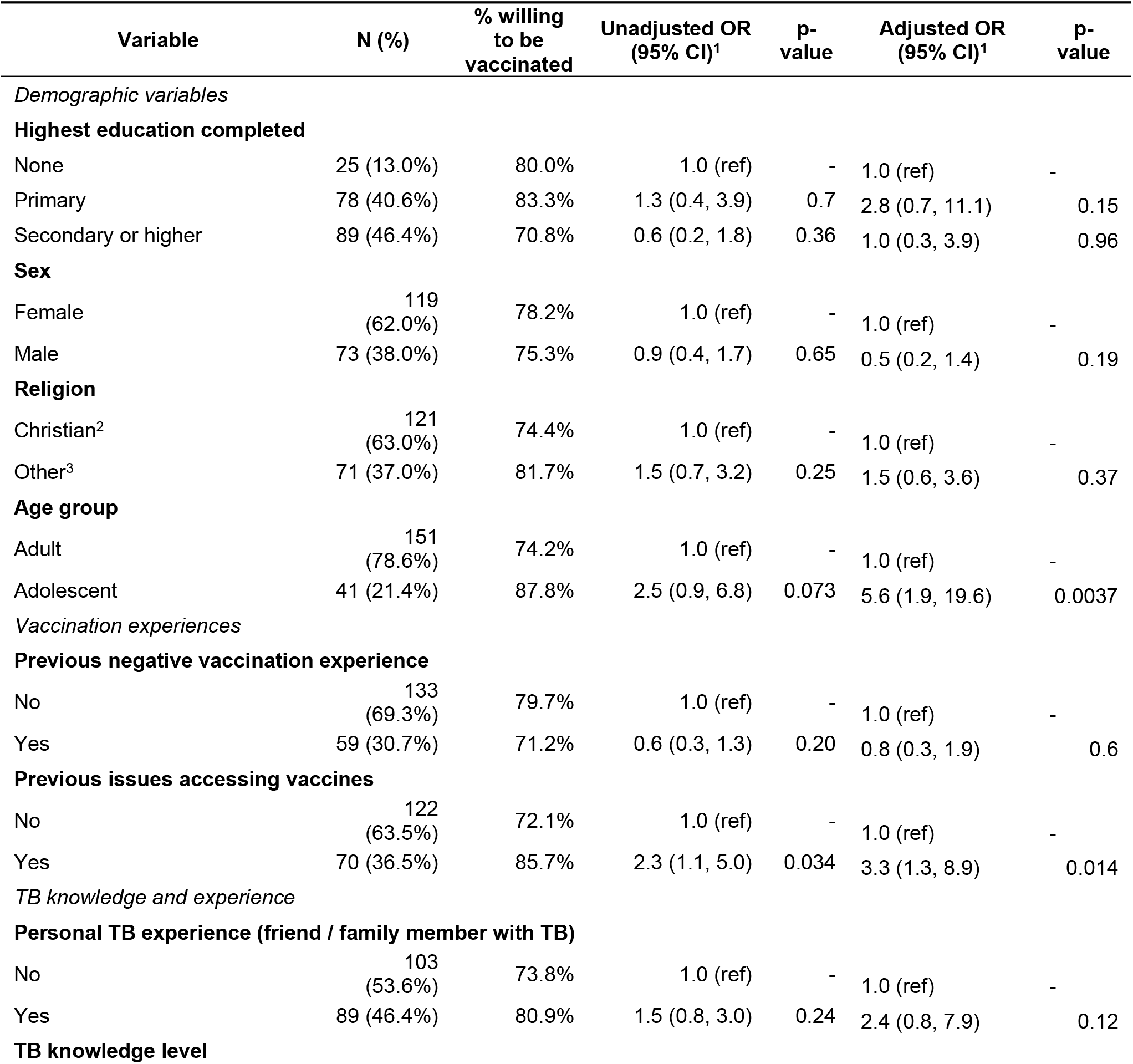
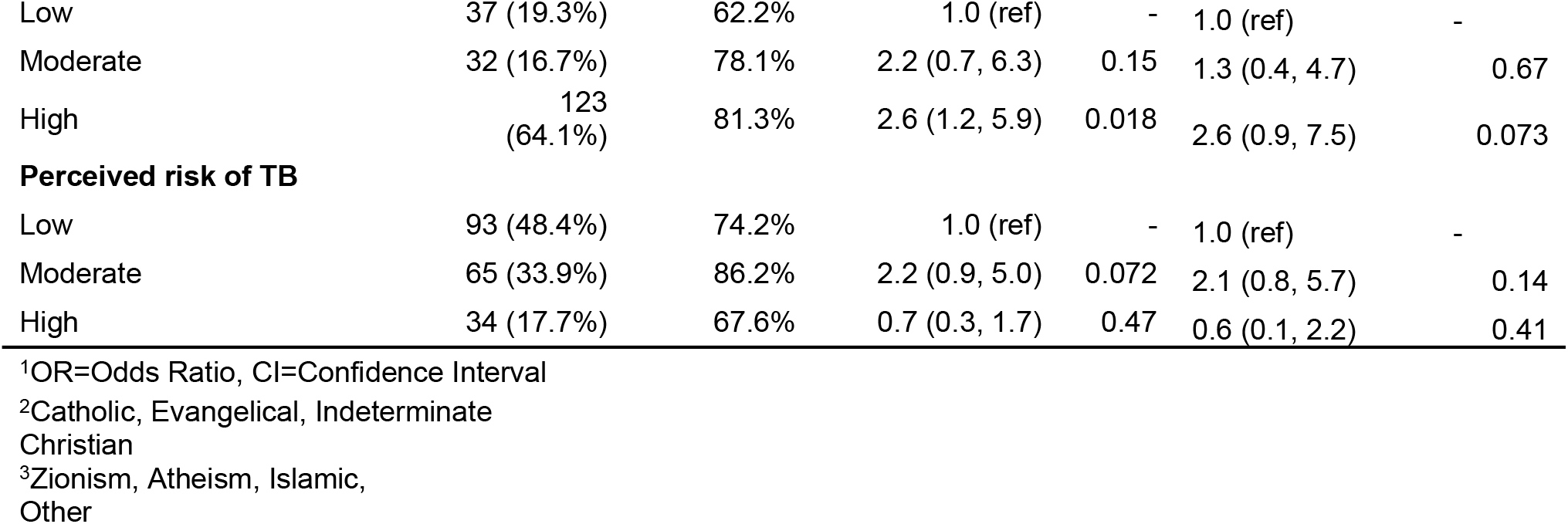
Associations between adult/adolescent characteristics and willingness to receive a novel TB vaccine in southern Mozambique, 2023-2024.

**Table 3.** Joint display of quantitative (survey) and qualitative (interview) findings.

| Finding from quantitative analysis [surveys] | Themes from qualitative analysis and illustrative quotes [interviews] | Integration of quantitative and qualitative findings |
| --- | --- | --- |
| **BESD domain: *Thinking and Feeling*** |  |  |
| Adolescents were more willing to receive a new TB vaccine than adults. (Source: Table 2) | Caregivers and adults said they wanted more information on why the vaccine would be offered to them and how it works.<br>Quote: "I am not saying I would not choose it, I do not know its behavior. I do not know what it does after application, how it reacts in the person's body after application."-Adult | Surveys showed adults and caregivers were less willing to be vaccinated; interviews suggested this hesitancy was due to a need for more information on the vaccine. |
| Participants with a higher knowledge of TB were more willing to receive a new TB vaccine than those with lower knowledge. (Source: Table 2) | Participants had a general understanding of TB as an illness (i.e. symptoms), but various misconceptions around transmission highlighted some knowledge gaps. Some cited health behaviors (i.e. unprotected sex or alcohol use) as leading to *Mtb* infection.<br>Quote: "as I understand it, I can transmit it to another person when, for example, I cough without covering my mouth, and my saliva passes to the other person" - Adult | Survey data demonstrated statistical associations between knowledge of TB and willingness to vaccinate; interviews revealed the types of misconceptions held about TB. |
| Participants reporting personal experience with TB (a friend or family member with the disease) were more willing to receive a new TB vaccine than those who did not. (Source: Table 2) | Participants who knew someone with TB expressed the importance of preventing TB through all means (e.g., treatment of *Mtb* infection, regular health screenings). Vaccination was seen as another method of prevention that would be important for protecting themselves and other family members, especially young children.<br>Quote: "The vaccine will prevent the disease I have and yes, I will feel good about my health." -Adolescent with TB | Surveys demonstrated an association between personal experience with TB and willingness to receive a vaccine. Interviews showed that participants with experience of TB felt that all prevention methods were important but especially saw value in a vaccine to keep the community safe. There was great emphasis on making sure other family members (particularly young children) were safe from *Mtb* infection, which was explained in in-depth interviews but not explored in the survey. |
| Not assessed. | Only a few participants expressed true doubt or hesitancy. Concerns focused on vaccine development and testing in a location not deemed 'technologically advanced'. | - |
| **BESD domain: *Social Processes*** |  |  |
| Most trusted sources of information were healthcare providers, government agencies; participants had relatively little trust in information from social networking sites. (Source: Supplemental Table 1) | Information sources included advertisements, social media, and outreach workers. Many expressed distrust of messages via social platforms (i.e. WhatsApp). | Both surveys and interviews captured similar data on sources of trusted information: participants reported that messages from the health system were trusted and information coming from social networking platforms was not. |
|  | Quote: "...but if I have any doubts, I should go to a health center and tell them that I heard that information on television, so I went there to find out if it was true, and then they'll clarify it for me." – Adult |  |
| People most influencing a person's decision to receive a vaccine were health providers/community health workers, themselves (the respondents), and local community leaders. (Source: Supplemental Table 2) | Social influences are described as significant motivators. These include healthcare professionals, the neighborhood 'secretary', family, community leaders, and spouses.<br>Quote: "I chose these people because they know what they're talking about, they can't mislead me." – Caregiver | Both surveys and interviews captured similar data on who influences vaccination decisions, showing a strong influence of other family members, healthcare providers, and community leaders. |
| **BESD domain: *Motivation*** |  |  |
| Participants who reported previous issues accessing vaccines were more willing to receive a new TB vaccine than those who did not. (Source: Table 2) | Prior vaccination was reported as generally a positive experience, even if minor side effects were felt.<br>Quote: "When I vaccinated at school, I felt good because it was necessary for me to be vaccinated for the good of my health but nothing went wrong with me, I liked the vaccine" - Adult | Surveys showed that previous issues accessing vaccines made individuals more willing to receive a TB vaccine. This was discordant with interview data, in which a majority of respondents reported that previously receiving vaccines and having positive experiences with accessing and receiving them led to an interest in the new vaccine. |
| Most important factors cited in influencing vaccination decisions were whether the vaccine prevented disease and was deemed 'important'. (Source: Supplemental Table 3) | Participants felt that if they were assured the vaccine could prevent *Mtb* infection and keep them healthy, they would be willing to take it.<br>Quote: "so that you can be vaccinated and you'll have a lot of health in your life..." - Adult | Both surveys and interviews showed that effectiveness of the vaccine in preventing *Mtb* infection/TB is an important factor in vaccination decisions. Survey data additionally suggested that participants also valued a healthcare professional's evaluation of whether it was 'important' for their health. |
| **BESD domain: *Practical Issues*** |  |  |
| Not assessed. | Locations like schools, clinics, and hospitals that serve as education centers were also seen as possible places of vaccine administration.<br>Quote: "The advantage of the school is that everything they teach there comes from the Ministry of Health, so I trust the school." - Adult | - |

### Interview findings

We recruited 23 survey respondents (6 adolescents, 11 adults, and 6 caregivers) to participate in in-depth interviews. Nineteen (n=19, 79%) interviewees reported in the survey that they would receive a TB vaccine, four (n=4, 21%) said they would not. Due to the sparseness of qualitative data from some groups, especially adolescents who gave brief answers to interview questions and the overall low number of participants who responded they would not receive a TB vaccine, we present below the combined qualitative and quantitative findings of our study.

### Integration of qualitative and quantitative findings by BeSD domain

#### Thinking and Feeling

Sentiments expressed in interviews generally substantiated the quantitative findings from the survey. Consistent with the finding that adults were less willing than adolescents to receive a new TB vaccine, adults expressed the need for more information on why the vaccine is being offered to them and how it will work before receiving, or allowing their child to receive, a new TB vaccine. Persons with greater knowledge of or experience with TB were more willing to receive a TB vaccine, and findings from the interviews were largely consistent with this finding. Specifically, interviewees expounded upon some misconceptions held about TB but also the importance placed on prevention measures that would protect vulnerable community members such as children.

#### Social Processes

Surveys and interviews provided consistent information on how vaccination decisions are made and the most influential sources of information. In the survey, respondents identified healthcare professional, government agencies, and community leaders as trusted sources of information that would influence vaccination decisions (Supplemental Tables 1-2). In interviews, respondents identified healthcare providers and government agencies as reliable sources of information about a TB vaccine. Community health workers and community leaders were identified as important influences on their vaccination decisions; social media was usually cited as a less trusted source of information.

#### Motivation

In the survey, people who reported previous issues accessing vaccines were *more* willing to receive a new TB vaccine (Table 2). In interviews, there were few past negative vaccination experiences influencing attitudes towards a new TB vaccine; in contrast, most previous vaccine experiences were positive even if minor side effects were noted. While superficially discordant, this may reflect generally positive attitudes towards vaccines, such that even people who may have had issues accessing vaccines in the past were not deterred from receiving a new TB vaccine. In interviews, participants explained that if they could be assured the vaccine could effectively prevent TB, they would be willing to receive it.

#### Practical Issues

Finally, interview responses revealed a preference for vaccination at schools, clinics, and hospitals, where people are already accustomed to both receiving health information and interventions.

## Discussion

We conducted a mixed methods study on willingness to receive a new TB vaccine among the target population for new vaccines (i.e., adults and adolescents) and caregivers of adolescents in southern Mozambique. We found an overall high proportion of willingness to receive a new TB vaccine (77%), which anticipates moderate to high uptake of a TB vaccine in this setting.^17^ We now report that adolescents were more willing to receive a new TB vaccine than adults even when adjusting for other factors which may influence vaccination decisions. Through in-depth interviews which further explored the preferences expressed in survey responses and largely corroborated the quantitative findings from surveys, we found that participants expressing uncertainty about vaccination would require more information about how a new vaccine would work and why it is being recommended to them. However, most felt that if a vaccine was shown to prevent TB, they would be willing to receive it. Healthcare workers, government agencies, and community leaders were cited as trusted voices of information about a new TB vaccine, with social media as relatively less important in influencing vaccination decisions.

This is the first study of which we are aware to explore community perceptions of new TB vaccines, so there is little prior data on this topic to which to compare our findings. However, we note that acceptability and uptake of another preventive intervention against TB, drug therapy to prevent the progression of *Mtb* infection to TB (i.e., TB preventive therapy, or TPT), is generally low compared to our estimates of willingness to receive a TB vaccine. Estimates of TPT uptake in South Africa are approximately 50% among persons in whom it is indicated, compared to 77% of respondents in our study who responded that they would receive a new TB vaccine if available.^18^ If a TB vaccine for adolescents and adults is approved, understanding different attitudes towards these two preventive interventions can set expectations about the uptake of each in different settings and population groups. Despite apparently higher acceptability than TPT, willingness to receive a TB vaccine in our study was lower than for other new vaccines. A systematic review of malaria vaccine acceptance studies found 95% acceptability in surveys completed prior to vaccine introduction in malaria-endemic countries.^19^

We found several factors associated with willingness to be vaccinated. Willingness to be vaccinated was associated with greater knowledge of TB, suggesting that a basic level of knowledge about the disease is key to understanding the need for a vaccine. Relatedly, persons who knew someone with TB were also more willing to be vaccinated and, in interviews, expressed the importance of preventive interventions against TB. These findings suggest that general health education about TB (on its symptoms, treatment, and transmission) in conjunction with information about a new vaccine could facilitate high uptake in this setting. Adults were generally more skeptical of vaccines than were adolescents, especially adult men. Given the higher burden of TB among adult men, and their disproportionate contribution to *Mtb* transmission, vaccine outreach will be a particularly important for this group. Women expressed more uncertainty about vaccination than men, expressing a need for more information prior to making a decision about vaccination, again underscoring the need to clearly communicate about the key features of and need for a new adolescent and adult TB vaccine, given that there is an existing vaccine for TB (BCG). Additional studies should continue to characterize reasons for TB vaccine hesitancy that may result in lower levels of acceptance for a new TB vaccine to understand ways to remove barriers to vaccine uptake and to design tailored communication strategies that address specific community concerns about vaccination. In this area of southern Mozambique, we found that healthcare providers, government agencies (i.e., the Ministry of Health), and community leaders were trusted sources of vaccine information. Provider recommendation is a well-documented strategy for increasing uptake of other vaccines^20^ and may be an important strategy for maximizing uptake of new TB vaccines. Social media was not considered a trusted source of information, though the reach of these platforms is still low in Mozambique (<12% of the population in early 2025 uses Facebook).^21^ Understanding the most credible sources of vaccine information can guide decisions about the platforms of communication campaigns suitable for different settings and population subgroups. Work to understand optimal communication strategies will be particularly important given that there are few other vaccines in Mozambique with broad indications in adolescents and adults; the methods for effectively reaching this group may be different from those typically used to encourage childhood vaccination. Recent immunization campaigns in this age group for HPV and SARS-CoV-2 vaccines have been successful^22,23^ and may be a useful template for new TB vaccines.

There were two surprising findings that merit additional research. We found that adults who reported previous issues accessing vaccines were more likely to receive a new TB vaccine. While we think this may reflect high motivation to be vaccinated, combined with the recent memory of limited supply of SARS-CoV-2 vaccines in Mozambique, this dynamic should be explored in future studies. In addition, adults who reported high perceived risk of TB were less likely to be vaccinated than those who perceived themselves at moderate risk of TB. While these trends were not statistically significant, we consider this an area where additional qualitative work may explain these attitudes and offer further insights to facilitate vaccine uptake.

Our study has several limitations. First, this was a small study (n=241) recruited using purposive sampling. Our study population over-represents women (68%) and individuals with past TB (56%). It should not be considered representative of Mozambique nationally, or of other countries, since attitudes towards vaccines are known to vary significantly by setting. Second, we encountered difficulties with eliciting rich interview responses from adolescents, especially adolescent males. (Many responses were short in duration and lacking in depth.) We used the interview findings to substantiate and elaborate on results from the quantitative analysis rather than conduct a standalone qualitative analysis, in part because of the sparse responses given by adolescents. More work should be done among adolescents to understand attitudes towards a new TB vaccine, as this period represents a time of increasing TB exposure risk and therefore a key population group for ensuring high coverage of preventive interventions. Moreover, since we collected qualitative and quantitative data collection simultaneously, emergent findings in the qualitative data could not be explored quantitatively (and vice versa). Future mixed methods work will be useful to more fully explore attitudes around new TB vaccines in this setting. Finally, we acknowledge that willingness to be vaccinated may not necessarily translate into a decision to vaccinate, and therefore these estimates should not be interpreted as expectations of vaccine coverage. However, these data remain important for understanding potential uptake, anticipating barriers to high coverage in this setting, and suggesting avenues for vaccine outreach to plan for the eventual introduction of an adolescent and adult TB vaccine.

In summary, our study reports high willingness to receive a new TB vaccine among adolescents and adults, with adolescents especially willing to receive a new TB vaccine. Personal experience with TB and greater knowledge of the disease was linked with willingness to be vaccinated. Qualitative findings largely reinforced quantitative findings, further clarifying that even among those who expressed hesitancy, a safe and effective TB vaccine that is endorsed by healthcare workers, government agencies, and community leaders would likely have high uptake. Our findings are specific to southern Mozambique and can shape vaccine introduction efforts after a TB vaccine is licensed and approved for use in this age group.

## Data Availability

An abbreviated dataset and the scripts used for the descriptive and multivariable analysis of survey data are available on GitHub (https://github.com/kbratnelson/tbvax-willingness-moz). Qualitative data is available upon reasonable request subject to ethical approval and data privacy restrictions.

## Declarations

### Ethics approval and consent to participate

This study was approved by ethical and scientific committees at Centro de Investigação em Saúde da Manhiça (CISM), Manhiça, Mozambique (CIBS-CISM/050/2023 and CCI/036/AGO/2023) and Emory University (study #00006480). All study procedures were developed and carried out in accordance with the Declaration of Helsinki. Written consent was obtained from all participants over the age of 17. For participants aged 17 and under, written assent and parental consent were obtained. Clinical trial number: not applicable.

### Consent for publication

Not applicable.

### Authors’ contributions

KNN, LV, LMC, and AL designed the study. KNN obtained funding for the study. AL, SA, and AGB led data collection. KNN, IC, DK, and MS conducted data analysis. KNN drafted the manuscript. All authors critically reviewed the manuscript and approved the final version.

## Acknowledgements

We are grateful for the time and effort contributed by the participants. We thank Neusa Torres and the social sciences team, data managers, field staff at the Manhiça Health Research Center, and Casey Randleman and Shamika Chavda for project and analysis support at Emory University.

